# The genetic analysis of a founder Northern American population of European descent identifies *FANCI* as a candidate familial ovarian cancer risk gene

**DOI:** 10.1101/2020.05.04.20090407

**Authors:** Caitlin T Fierheller, Laure Guitton-Sert, Wejdan M Alenezi, Timothée Revil, Kathleen K Oros, Karine Bedard, Suzanna L Arcand, Corinne Serruya, Supriya Behl, Liliane Meunier, Hubert Fleury, Eleanor Fewings, Deepak N Subramanian, Javad Nadaf, Diane Provencher, William D Foulkes, Zaki El Haffaf, Anne-Marie Mes-Masson, Jacek Majewski, Marc Tischkowitz, Paul A James, Ian G Campbell, Celia M T Greenwood, Jiannis Ragoussis, Jean-Yves Masson, Patricia N Tonin

## Abstract

Some familial ovarian cancer (OC) could be due to rare risk alleles in genes that each account for a relatively small proportion of cases not due to *BRCA1* and *BRCA2*, major risk genes in the homologous recombination (HR) DNA repair pathway. We report a new candidate OC risk allele, *FANCI* c.1813C>T in a Fanconi anemia (FA) gene that plays a role upstream of the HR DNA repair pathway. This variant was identified by whole exome sequencing of a *BRCA1* and *BRCA2* mutation-negative French Canadian (FC) OC family from a population exhibiting founder effects. In FCs, the c.1813C>T allele was detected in 7% (3/43) of familial and 1.6% (7/439) of sporadic OC cases; and in 3.7% (3/82) of familial breast cancer (BC) cases with a family history of OC and in 1.9% (3/158) of BC only families. This allele was significantly associated with FC *BRCA1* and *BRCA2* mutation-negative OC families (OR=5.6; 95%CI=1.6-19; p=0.006). Although *FANCI* c.1813C>T was detected in 2.5% (74/2950) of cancer-free FC females, carriers had a personal history of known OC risk reducing factors, and female/male carriers were more likely to have reported a first-degree relative with OC (ρ=0.037; p=0.011). Eight rare potentially pathogenic *FANCI* variants were identified in 3.3% (17/516) of Australian OC cases, including 10 carriers of *FANCI* c.1813C>T. Potentially pathogenic *FANCI* variants were significantly more common in AUS OC cases with a family history of OC than in isolated OC cases (p=0.027). The odds ratios (OR) were >3 for carriers of any of the seven rarest *FANCI* alleles, and 1.5 for c.1813C>T. Data from the OC Association Consortium revealed that the ORs for the c.1813C>T allele were highest for the most common OC subtypes. Localization of FANCD2, part of the FANCI-FANCD2 (ID2) binding complex in the FA pathway, to sites of induced DNA damage was severely impeded in cells expressing the p.L605F isoform. This isoform was expressed at a reduced level; unstable by formaldehyde or mitomycin C treatment; and exhibited sensitivity to cisplatin but not to olaparib (a poly [ADP-ribose] polymerase inhibitor). By tissue microarray analyses, FANCI protein was robustly expressed in fallopian tube epithelial cells but expressed at low-to-moderate levels in 88% (83/94) of high-grade serous carcinoma OC samples. This is the first study to describe potentially pathogenic variants in OC in a member of the ID2 complex of the FA DNA repair pathway. Our data suggest that potentially pathogenic *FANCI* variants may modify OC risk in cancer families.

---

Ovarian cancer (OC), with an overall five-year survival rate of 40%, is the leading cause of death in women with gynecologic cancen. The overall lifetime risk for OC in the North American population is 1.3%^1^. However, twin studies suggest that 22% of OC risk can be attributed to heritable factors^2^ and having an affected first-degree relative confers a 3-7 fold increase in risk to this disease^3,4^. Carriers that are heterozygous for pathogenic variants in *BRCA1* (*FANCS*) or *BRCA2* (*FANCD1*) have an estimated lifetime risk for OC of 17-44% (by age 80 years), depending on the gene mutated^5^. Pathogenic *BRCA1* and *BRCA2* variants have been reported in 65-85% of cancer syndromes featuring high-grade serous carcinoma (HGSC)^6^ - the most common histopathological subtype of epithelial OC^7^ - and/or young age (<50 years) of onset breast cancer (BC), and in 10-20% of HGSC cases regardless of age at diagnosis^8^. Identifying carriers of *BRCA1* and *BRCA2* pathogenic variants for cancer prevention (prophylactic surgery^9,10^) and increasingly, management of OC using new therapies (poly [ADP-ribose] polymerase inhibitors [PARPi]^11-16^) has been offered in medical genetic and gynecologic oncology settings.

New cancer predisposing gene (CPG) candidates have been investigated with a focus on members of the Fanconi anemia (FA) DNA repair pathway involving BRCA1 and BRCA2 function. An early study of 429 women with HGSC reported that 20% are carriers of heterozygous loss of function (LoF) variants in FA pathway genes, though their association with heritable risk was not investigated^17^. The most promising new OC predisposing genes are from reports of heterozygous carriers of potentially pathogenic variants in *BRIP1* (*FANCJ*)^18,19^, *RAD51C* (*FANCO*)^20-23^, and *RAD51D*^24^. In cancer families carriers of pathogenic *RAD51C* and *RAD51D* variants have been estimated to have cumulative risks to age 80 of 11% (95%CI 6-21) and 13% (95%CI 7-23), respectively, for OC^25^. Collectively, carriers of pathogenic variants in these genes have not accounted for a large proportion of familial OC and BC cases that have not been attributed to the known CPGs. Therefore, it is possible that new CPGs conferring risk to OC have yet to be discovered.

The low incidence of OC, rarity of pathogenic variants in each proposed CPG candidate, and genetic heterogeneity of the general population pose major challenges in finding new OC predisposing genes. An attractive strategy for gene discovery focuses on the investigation of demographically (ethnically/geographically) defined populations that take advantage of founder effects. Due to common ancestry, rapid expansion and geographic isolation during 1608~1760 of the small founding immigrant French population of Quebec from Europe (EUR), a loss of genetic variation has occurred resulting in subsequent waves of expansion of specific variants^26-29^. As French Canadians (FC) are more likely to harbour recurrent germline pathogenic variants, candidate variants are readily identified by sequencing familial cases and/or by comparing allele frequencies in cancer cases versus cancer-free controls in contrast to studies involving the general population due to allelic heterogeneity^26,27^. Though 19 different pathogenic *BRCA1* or *BRCA2* variants have been identified in FC cancer families of Quebec, five recurrent pathogenic variants account for 84% of all mutation-positive BC and/or OC families^30^. This is in contrast to the over two thousand different pathogenic *BRCA1* and *BRCA2* variants reported for undefined populations^31^. Specific recurrent pathogenic variants in *PALB2* (*FANCN* - c.2323C>T; p.Q775X)^32^ and *RAD51D* (c.620C>T; p.S207L)^33^ have since been identified in BC and HGSC cases of FC descent, respectively.

Using whole exome sequencing (WES) analyses, we identified heterozygous carriers of *FANCI* c.1813C>T; p.L605F missense variant among OC affected sisters in a *BRCA1* and *BRCA2* mutation-negative four-case OC family of FC descent. We pursued this variant as a candidate OC risk allele as it occurred in the FA Complementation Group I gene which is purported to play a role upstream of the homologous recombination (HR) DNA repair pathway involving BRCA1 and BRCA2 function^34-36^. Also, FANCI is an essential member of the FA-HR DNA repair pathway and acts as the molecular switch to activate this pathway^37^. In response to DNA damage, FANCI and its heterodimeric binding partner FANCD2, which form the ID2 complex, are monoubiquitinated and localized to sites of DNA damage allowing for the recruitment of other DNA repair proteins^34-36,38^.

To evaluate the potential pathogenicity of *FANCI* c.1813C>T; p.L605F, we applied a genetic strategy that took advantage of founder effects in the FC population by investigating its allele frequency in FC OC and cancer-free subjects. We applied various *in vitro* experiments to investigate the effects of the encoded p.L605F isoform on FANCI protein function and response to chemotherapies used in the treatment of OC. We also investigated FANCI expression in HGSC and normal tissues. Then, based on our findings from these studies, we investigated other populations, specifically Canadian non-FC (CDN) and Australia (AUS) familial cancer cases for rare potentially pathogenic *FANCI* variants.

## Results

### Discovery of *FANCI* c.1813C>T as a candidate OC predisposing variant

We previously reported a rare *BRCA1* and *BRCA2* mutation-negative four-case OC family (F1528) in a study of the histopathology of OC and *BRCA1* and *BRCA2* pathogenic variant carrier status of FC cancer families^39^. To investigate if other potentially pathogenic variants could be contributing to cancer risk in this family, WES and bioinformatic analyses were performed on peripheral blood lymphocyte (PBL) DNA available from two affected siblings both of whom had HGSC^39^. We selected rare (variant allele frequency [VAF] <1%) potentially pathogenic variants as candidates that were inherited in the heterozygous state and shared in common with the affected sisters. *FANCI* c.1813C>T was among the list of rare variants shared by the affected sisters. This was an intriguing candidate to investigate given that family F1528 is predicted to harbour a pathogenic variant in *BRCA1* or *BRCA2* (Manchester score^40,41^: *BRCA1=29, BRCA2=20*), genes which are involved in the FA-HR pathway. *FANCI*, which plays a role in this pathway, may be associated with phenotypically similar cancer families^39,42^ (**Figure 1**). The C>T nucleotide alteration is predicted to result in a leucine to phenylalanine change affecting a highly conserved amino acid residue at position 605 (Genomic Evolutionary Rate Profiling [GERP++]^43^ score = 6.1) that resides in the S/TQ phosphorylation cluster^37^. It is also predicted to be “probably damaging” (PolyPhen2^44^ score = 1.0). However at the time of discovery, the overall allele frequency of *FANCI* c.1813C>T from available databases was 0.76% in the National Heart, Lung and Blood Institute (NHLBI) Exome Sequencing Project (ESP) v.2014 (https://evs.gs.washington.edu/EVS/) and 0.2% in the 1000 Genomes Project^45^. These allele frequencies were notably higher than expected for individual pathogenic variants found in known OC predisposing genes, such as *BRCA1* and *BRCA2* (0.001%). Therefore, we performed a molecular investigation before pursuing extensive genetic analyses of our study groups.

**Figure 1.**
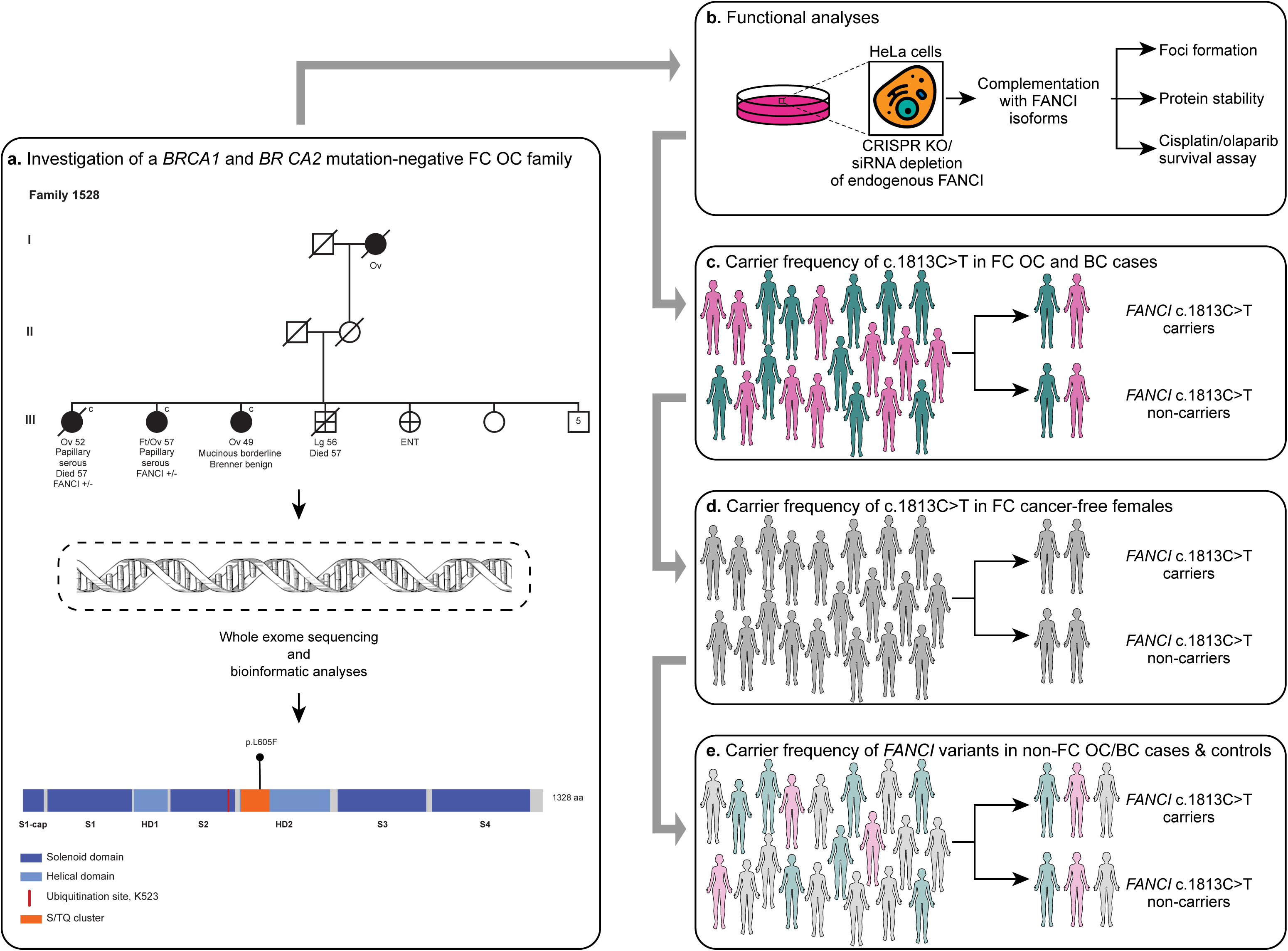
Study design for investigation of *FANCI* variants. A) Pedigree of a rare four-case FC OC family F1528 where *FANCI* c.1813C>T; p.L605F was discovered (top). Cancer type (ovarian [Ov], fallopian tube [Ft], lung [Lg], and ear, nose, and throat [ENT]) and age of diagnosis are shown. The affected Oc cases in generation III are *BRCA1* and *BRCA2* mutation-negative. WES was performed on PBL DNA from sisters (Ov 52 and Ft/Ov 57) from F1528 (middle). Protein location of p.L605F on FANCI (previously *KIAA1794;* bottom). B) Functional analyses of FANCI isoforms using HeLa cells. C-E) Investigation of carrier frequency of c.1813C>T in FC OC and BC cases (c) and cancer-free controls from CARTaGENE (d), and *FANCI* variants in non-FC OC/BC cases and controls. Solenoid domain=antiparallel pairs of α-helices that form α-α superhelix segments; Helical domain=α-helices; Ubiquitination site, K523=site of monoubiquitination by the FA core complex to allow downstream FA pathway function^48,49^; S/TQ cluster=location of conserved phosphorylation sites^51^

### *In vitro* analysis revealed FANCI p.L605F isoform behaves differently than wild-type protein

Using established *in vitro* assays, we observed that FANCI p.L605F was expressed as a full-length protein with severely affected capacity to localize to sites of DNA damage as compared with wild-type control. To further investigate the functional impact of FANCI p.L605F isoform, both HeLa CRISPR FANCI knock-out **(Figure 2a-g, Supplementary Figure 1)** or HeLa FANCI siRNA knockdown cells **(Supplementary Figure 1)** were complemented with the FANCI p.L605F isoform. Western blot analysis of cells treated with the DNA damaging agent mitomycin C (MMC) showed decreased levels of FANCI p.L605F isoform, unlike the FANCI p.P55L isoform encoded by polymorphic variant c.164C>T which has been reported to exhibit wild-type function^35^ **(Figure 2a, Supplementary Figure 1)**. Increasing the quantity of transfected *FANCI* c.1813C>T DNA by three-fold did not overtly affect the level of protein expression comparable to that seen in the wild-type FANCI or p.P55L isoform **(Supplementary Figure 1)**. Essential to downstream FA pathway function is the interdependent monoubiquitination of both FANCI and FANCD2^34-36^. Though FANCI p.L605F isoform co-immunoprecipitates with FANCD2, ubiquitination levels of FANCD2 were severely diminished as compared to those in FANCI wild-type expressing cells **(Figure 2a-b)**. These results are consistent with the decrease in the intensity of FANCD2 (see upper band in **Figure 2a**) after MMC treatment in cells complemented with FANCI p.L605F as compared to the WT or p.P55L isoforms. These observations suggest that while physical interactions between FANCI p.L605F isoform and FANCD2 proteins are maintained the altered FANCI isoform may affect monoubiquitination of FANCD2. Monoubiquitination of FANCD2 is required to form MMC-induced foci and consistent with this role the expression of FANCI p.L605F led to a significant reduction in the number of FANCD2 foci (**Figure 2c, Supplementary Figure 1**).

**Figure 2.**
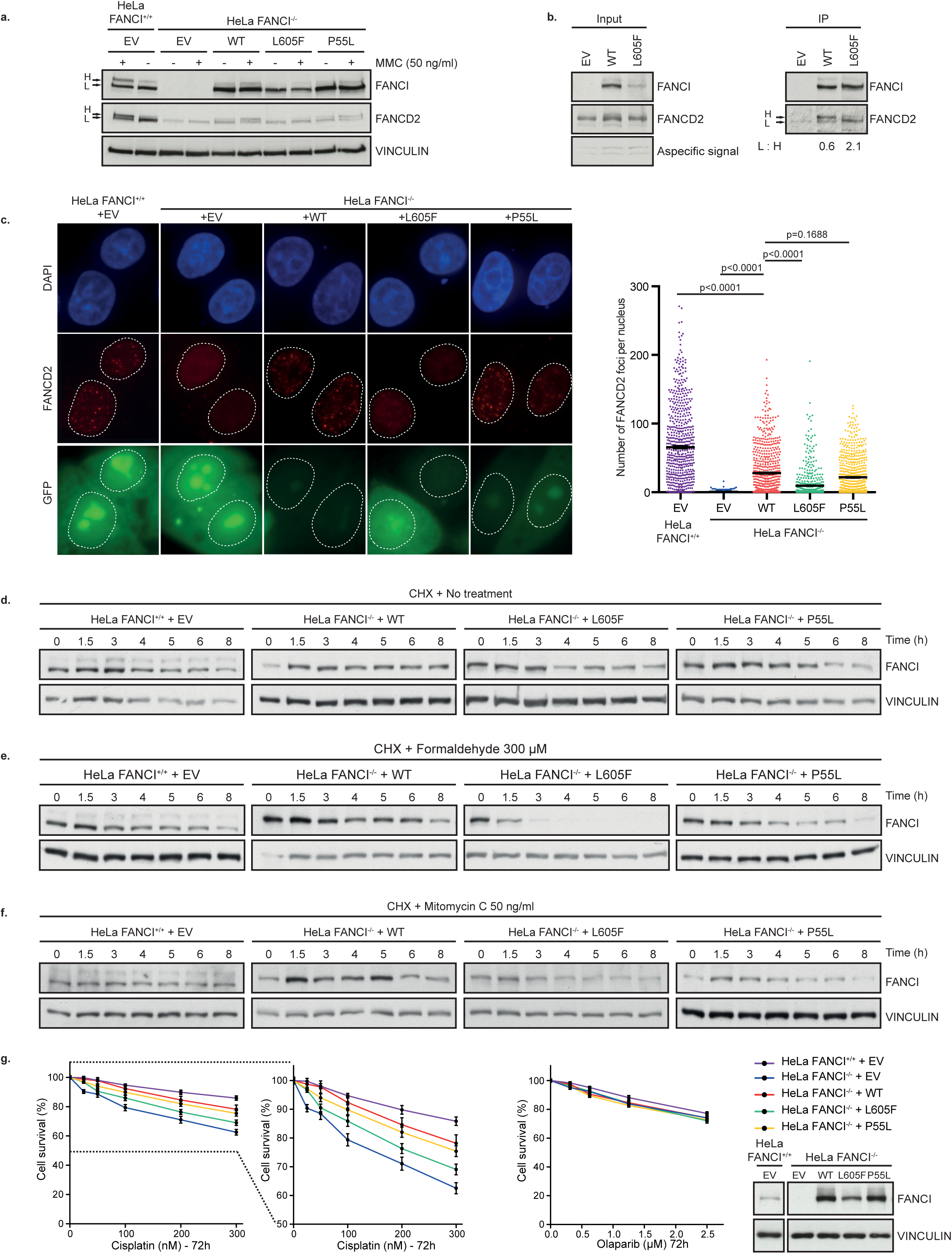
p.L605F variant impairs FANCI stability and functions. A) HeLa FANCI^-/-^ cells, clone 1, were complemented with indicated constructs of Flag-FANCI variants or empty vector (EV) and treated with 50 ng/ml MMC for 18 hours. Ubiquitination of FANCI and FANCD2 (upper band, H) after treatment was assessed by western blot. L=lower band, corresponds to non-ubiquitinated FANCI or FANCD2. B) HeLa FANCI^+/+^ cells were transfected with siRNA targeting FANCI and then complemented with Flag-FANCI siRNA resistant constructs or EV. Cells were treated with 50 ng/ml MMC for 18 hours followed by FLAG immunoprecipitation. Immunoprecipitated FANCD2 was analyzed by western blot. The ratio between the upper band (H) and lower band (L) was calculated. C) HeLa FANCI^-/-^ cells, clone 1, were complemented with indicated constructs of Flag-FANCI variants or EV and 0.1 µg of empty GFP vector. After treatment with MMC (50 ng/ml, 18 hours), FANCD2 foci were counted in GFP positive cells. Mean with SEM are shown, and Kruskall-Wallis test was performed. P-values are shown. D-F) HeLa FANCI-/- cells, clone 1, were complemented with indicated constructs and treated with CHX and either mock-treated (d) or treated with damaging agents (e, f) for indicated times. At each time point, whole cell extracts were analyzed by western blot to assess protein levels. G. HeLa FANCI^-/-^ cells, clone 1, were transfected with EV or indicated Flag-FANCI constructs. Cell viability was monitored following cisplatin or Olaparib treatments for 72 hours. Cell viability was assessed by counting remaining nuclei. Expression of Flag-FANCI constructs in HeLa FANCI^-/-^ complemented cells was monitored by western blotting.

As the expression of FANCI p.L605F appeared to be lower than the WT or p.P55L isoforms, even when increasing the quantity of plasmid **(Figure 2a, Supplementary Figure 1)**, we suspected that this protein isoform was unstable. To investigate protein stability, cells expressing FANCI wild type protein, or either of the p.L605F and p.P55L isoforms, were treated with cycloheximide (CHX) to inhibit protein synthesis and then treated with MMC or formaldehyde to induce DNA damage. FANCI protein levels decreased over time in response to both DNA damaging agents **(Figure 2d-f, Supplementary Figure 1)**. This effect was more prominent in FANCI p.L605F expressing cells as compared to wild-type FANCI or p.P55L expressing cells. These observations suggest that treatment with genotoxic agents exacerbates FANCI p.L605F protein instability. This is in agreement with the observation that FANCI p.L605F failed to complement survival of the HeLa FANCI^-/-^ cells that were challenged with the platinum compound cisplatin **(Figure 2g, Supplementary Figure 1)**. In contrast, FANCI^-/-^ cells were not sensitive to olaparib, a PARPi **(Figure 2g, Supplementary Figure 1)**.

### *FANCI* c.1813C>T carriers are enriched in familial OC cases of FC descent

Based on the anticipated founder effects of the FC population^30,46,47^, we assessed *FANCI* c.1813C>T carrier frequency in available PBL DNA from index OC or BC cases of FC descent to determine if this variant plays a role in conferring risk in phenotypically defined cancer families^18,19,48-55,20,21,23,24,30,32,33,39^. These OC or BC cases were selected based on their family history of OC or BC, or regardless of cancer family history (referred to as sporadic cases in this study), where *BRCA1* and *BRCA2* pathogenic variant carrier status was known^30,32,33,46-51,56-59^. Index OC cases from OC families (3/43, 7%) and index BC cases from hereditary breast and OC (HBOC) families (3/82, 3.7%) had a higher carrier frequency than BC cases from hereditary BC (HBC) families (3/158, 1.9%), though this difference was not statistically significant (p=0.41) **(Table 1, Figure 3, Supplementary Figure 2)**.

**Table 1.** Comparison of *FANCI* c.1813C>T carrier frequencies in cancer cases to cancer-free women of FC descent.

| Study group<sup>1</sup> | *BRCA1* and *BRCA2* mutation status<sup>1</sup> | Case tested | Number of subjects | Number of c.1813C>T carriers (%) | OR | 95% CI | P |
| --- | --- | --- | --- | --- | --- | --- | --- |
| OC families<sup>2</sup> | All | OC | 43 | 3 (7.0) | 2.91 | 0.88-9.64 | 0.08 |
|  | Negative |  | 23 | 2 (8.7) | 3.7 | 0.85-16.1 | 0.08 |
|  | *BRCA1* positive |  | 14 | 1 (7.1) | 2.99 | 0.39-23.2 | 0.29 |
|  | *BRCA2* positive |  | 6 | 0 | NA | NA | NA |
| Sporadic OC cases | All | OC | 439 | 7 (1.6) | 0.63 | 0.29-1.38 | 0.25 |
|  | Negative |  | 400 | 7 (1.8) | 0.69 | 0.32-1.51 | 0.36 |
|  | *BRCA1* positive |  | 18 | 0 | NA | NA | NA |
|  | *BRCA2* positive |  | 21 | 0 | NA | NA | NA |
| HGSC cases | All | OC | 341 | 7 (2.1) | 0.81 | 0.37-1.78 | 0.61 |
|  | Negative |  | 310 | 7 (2.3) | 0.9 | 0.41-1.97 | 0.79 |
|  | *BRCA1* positive |  | 15 | 0 | NA | NA | NA |
|  | *BRCA2* positive |  | 16 | 0 | NA | NA | NA |
| HBOC<sub>2</sub> | All | BC | 82 | 3 (3.7) | 1.48 | 0.46-4.78 | 0.52 |
|  | Negative |  | 34 | 2 (5.9) | 2.43 | 0.57-10.3 | 0.23 |
|  | *BRCA1* positive |  | 29 | 0 | NA | NA | NA |
|  | *BRCA2* positive |  | 21 | 1 (4.8) | 1.94 | 0.26-14.7 | 0.52 |
| HBC | All | BC | 158 | 3 (1.9) | 0.75 | 0.23-2.41 | 0.63 |
|  | Negative |  | 93 | 2 (2.2) | 0.85 | 0.21-3.53 | 0.83 |
|  | *BRCA1* positive |  | 20 | 1 (5) | 2.05 | 0.27-15.5 | 0.49 |
|  | *BRCA2* positive |  | 45 | 0 | NA | NA | NA |
| Sporadic BC cases | All | BC | 558 | 8 (1.4) | 0.57 | 0.27-1.18 | 0.13 |
|  | Negative |  | 538 | 8 (1.5) | 0.59 | 0.28-1.22 | 0.16 |
|  | *BRCA1* positive |  | 4 | 0 | NA | NA | NA |
|  | *BRCA2* positive |  | 17 | 0 | NA | NA | NA |
| Healthy women | NA | Na | 2950 | 74 (2.5) | NA | NA | NA |

<sup>1</sup>See Supplementary Table 1 for description of study groups
<sup>2</sup>Overlap of some families but not *FANCI* c.1813C>T carriers
NA – not available

**Figure 3.**
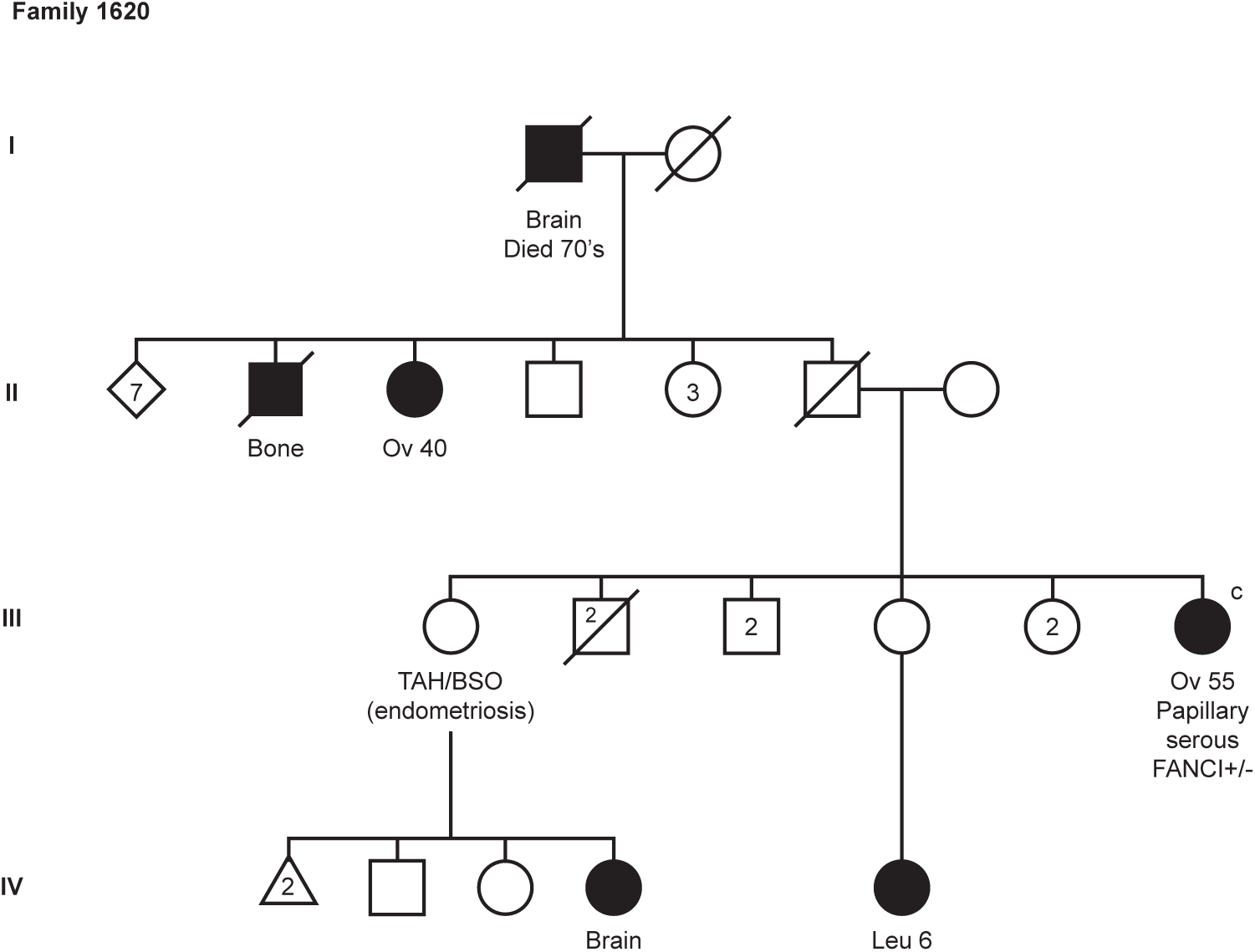
Pedigree of a *BRCA1* and *BRCA2* mutation-negative two-case FC OC family F1620 where *FANCI* c.1813C>T; p.L605F was identified by WES. Cancer type (ovarian [Ov] and leukemia [Leu]), risk reducing surgery (total abdominal hysterectomy/bilateral salpingo-oophorectomy [TAH/BSO]), and age of diagnosis are shown.

The *FANCI* variant carrier frequency was also higher in sporadic OC cases (7/439, 1.6%) than sporadic BC cases (8/558, 1.4%) though this difference was not statistically significant (p=1) (**Table 1**). When taken together, *FANCI* c.1813C>T variant carrier frequency was highest in index OC or BC cases from OC and HBOC families, respectively, and sporadic OC cases as compared with sporadic BC cases (**Table 1**). The highest frequency of *FANCI* variant carriers was in OC index cases from OC families (at 9.1% [all] or 12.5% [*BRCA1* and *BRCA2* mutation-negative]) when also including the multi-case discovery family in this group, which is significantly higher compared to sporadic OC cases (p=0.012 [all] or p=0.01 [*BRCA1* and *BRCA2* mutation-negative]).

### Cancer-free FC *FANCI* c.1813C>T carriers are significantly correlated with having a first-degree relative with OC

Since our initial discovery of the *FANCI* c.1813C>T variant, new data has become available from the CARTaGENE biobank enabling the evaluation of its allele frequency in study subjects from a cancer-free female FC population, and thus providing a more comparable reference group to our FC cancer subjects^60^. Using data from three different genotyping platforms, we estimated a 1.2% VAF in cancer-free FC female (**Supplementary Table 2**). This is not significantly different from the 1% estimated VAF in non-Finnish EURs, a population most likely to share common ancestry with FCs (France)^26,27^, as reported in the Genome Aggregation Database (gnomAD)^61^ (**Supplementary Table 2**). In this database the VAF varied across populations: highest in Estonians (2.1%) to none in East Asians.

The estimated heterozygous carrier frequency at 2.5% in cancer-free FC females was lower than that observed in index cancer cases from OC (7%) and HBOC (3.7%) families, but higher than observed in sporadic OC cases (1.6%) and index BC cases from HBC families (1.9%), though these differences were not statistically significant (**Table 1**). Notable was a trend towards statistical significance for the presence of variant carriers in OC families with the inclusion of the multi-case OC discovery family in our analysis (p=0.026 [all] or p=0.023 [*BRCA1* and *BRCA2* mutation-negative]).

Additional information was available from the CARTaGENE subjects to investigate *FANCI* c.1813C>T carrier frequency in the context of cancer family history (first-degree only), reproductive history, oral contraceptive pill (OCP) use, oophorectomy, and tubal (fallopian) ligation; all of which are factors that are known to significantly impact lifetime risk of OC^62,63^. We observed that cancer-free variant carriers (male or female) were significantly correlated with having a first-degree relative with OC (ρ=0.037; p=0.011), when analyzing data from subjects genotyped with arrays that included probes for the variant allele (n=7272) (**Supplementary Table 3**). This finding is consistent with OC family history as a risk factor for OC^4^. When adding data from cancer-free female and male subjects where genotypes were imputed (n=927) variant allele carriers were correlated with having a first-degree relative with OC or BC (ρ=0.026; p=0.046) (**Supplementary Table 3, Supplementary Table 4**). No other cancer type was significantly correlated with variant carrier status. Notably, the majority of cancer-free FC females were parous (78%; 2315/2950) and 91.8% (2710/2950) had experienced OCP use, oophorectomy, and/or tubal ligation (**Supplementary Table 5**), as all of these factors are known to significantly decrease OC risk^9,10,64-73^. Only 8.1% (6/74) of variant carriers reported having never experienced any of these risk reducing events.

### Other potentially pathogenic *FANCI* variants are rare in OC and BC cases of FC descent

To determine if there are other potentially pathogenic *FANCI* variants (VAF<1%) in FCs, we investigated available WES data from 81 familial and/or young age of onset OC cases and 76 BC cases from HBC/HBOC families, regardless of *BRCA1* or *BRCA2* pathogenic variant carrier status (**Supplementary Table 1**). In all, we identified seven rare variants among 33 index OC familial cases, where one carrier was heterozygous for *FANCI* c.1573A>G; p.M525V (**Supplementary Table 6**). Although the missense variant is predicted to result in a methionine to valine change affecting a highly conserved amino acid residue at position 525 (GERP++ score = 5.68) and to be potentially pathogenic by 7/14 *in silico* tools, our *in vitro* analyses suggested that it does not encode an aberrantly functioning protein (data not shown). Thus, *FANCI* c.1813C>T is the only plausible potentially pathogenic variant identified in FC OC/BC cases (**Figure 4a**).

**Figure 4.**
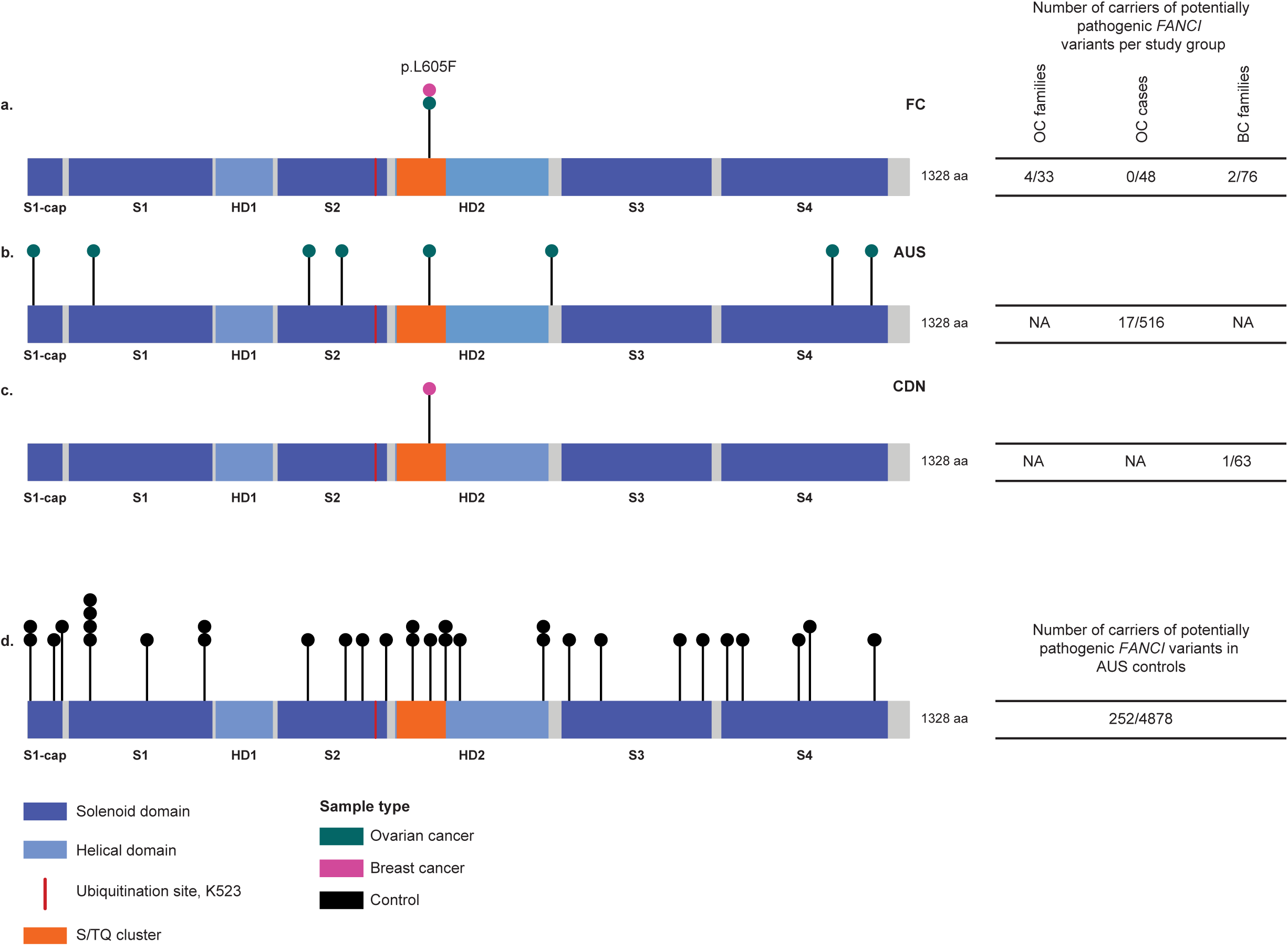
The location of potentially pathogenic variants in *FANCI* from different study groups. Rare (VAF<1%), potentially pathogenic *FANCI* variants in FC (a), AUS (b), CDN (c) OC and/or BC cases, and AUS controls (d). Refer to supplementary table 1 for cohort descriptions.

### OC and BC cases of non-FC descent also carry potentially pathogenic *FANCI* variants

To determine if there are carriers of rare potentially pathogenic *FANCI* variants in non-FC populations, we investigated available WES data from 516 AUS HGSC cases^74^ and 63 CDN BC families, all *BRCA1* and *BRCA2* mutation-negative (**Supplementary Table 1**). We also investigated available sequencing data for *FANCI* from 4,878 AUS cancer-free controls. Principal component analysis of the AUS cases and controls using data from ethnicity associated single nucleotide polymorphisms (SNP) suggested that >95% of these cases are of Western EUR ancestry, whereas mixed EUR ancestry was reported for CDN cases.

In all, we identified a total of 18 and 92 rare (VAF<1%) *FANCI* variants in the AUS HGSC cases and controls, respectively. There were 17 carriers of eight potentially pathogenic missense variants in 516 HGSC cases (3.3%), where 10 (1.9%) carried *FANCI* c.1813C>T (**Table 2, Figure 4b, d, Supplementary Table 6**). In AUS controls, we identified 133 carriers of 32 different potentially pathogenic missense variants (2.7%), where 95 (1.9%) carried *FANCI* c.1813C>T. The number of carriers of potentially pathogenic variants in *FANCI* was not significantly different between AUS cases and controls (p=0.48), including *FANCI* c.1813C>T (p=1).

**Table 2.** Carrier frequencies of potentially pathogenic variants in *FANCI* from AUS HGSC cases and controls. 1

| Study group<sup>1</sup> | Number of subjects (%) | Number of variant carriers<sup>3</sup> (%) |  |  |  |  |  |  |  | Total (%) |
| --- | --- | --- | --- | --- | --- | --- | --- | --- | --- | --- |
|  |  | c.13A>G<br>p.I5V | c.286G>A<br>p.E96K | c.1264G>A<br>p.G422R | c.1412C>G<br>p.P471R | c.1813C>T<br>p.L605F<sup>4</sup> | c.2366C>T<br>p.A789V<sup>4</sup> | c.3635T>C<br>p.F1212S | c.3812C>T<br>p.S1271F |  |
| HGSC | 516 (100) | 1 (0.2) | 1 (0.2) | 2 (0.4) | 1 (0.2) | 10 (1.9) | 1 (0.2) | 1 (0.2) | 1 (0.2) | 17 (3.3) |
| Controls | 4878 (100) | 0 | 5 (0.1) | 7 (0.1) | 0 | 95 (1.9) | 0 | 0 | 1 (0.02) | 108 (2.2) |
| Family history of HGSC cases<sup>2</sup> |  | 0 | 0 | 0 | 0 | 0 | 0 | 0 | 0 | 0 |
| ≥2 OC cases (no BC) | 7 (1) | 0 | 0 | 0 | 0 | 0 | 0 | 0 | 0 | 0 |
| 1 OC case (no BC) | 49 (10) | 0 | 0 | 0 | 0 | 0 | 0 | 0 | 0 | 0 |
| ≥2 BC cases (no OC) | 45 (9) | 0 | 0 | 0 | 0 | 0 | 0 | 0 | 0 | 0 |
| 1 BC case (no OC) | 125 (24) | 0 | 1 (0.8) | 0 | 0 | 1 (0.8) | 0 | 0 | 0 | 2 (1.6) |
| ≥2 OC case and BC cases | 42 (8) | 0 | 0 | 1 (2.4) | 1 (2.4) | 5 (12) | 1 (2.4) | 0 | 0 | 7 (17) |
| 1 OC case (no BC) | 248 (48) | 1 (0.4) | 0 | 1 (0.4) | 0 | 4 (1.6) | 0 | 1 (0.4) | 1 (0.4) | 8 (3.2) |

<sup>1</sup>See Supplementary Table 1 for description of study groups
<sup>2</sup>First-, second-, or third-degree relation for OC; first- and second-degree relation only for BC
<sup>3</sup>See Supplementary Table 6 for more information on variants
<sup>4</sup>Variants carried by the same case (see Supplementary Table 7)

*FANCI* carriers were diagnosed with OC between the ages of 31-82 years (**Supplementary Table 7**). Five of the c.1813C>T carriers had a family history of OC within third degree relatives (5/98, 5.1%), which was significantly higher than the carrier frequency of this variant in isolated cases of HGSC cases (5/418, 1.2%; p=0.025) (**Table 2**). In contrast, there was no significant difference in the carrier frequency of *FANCI* c.1813C>T in cases with a reported family history of OC or BC (6/262, 2.3%) than those without (4/254, 1.6%; p=0.75). *FANCI* c.1813C>T co-occurred with another potentially pathogenic missense variant, *FANCI* c.2366C>T; p.A789V, in a HGSC case with a family history of OC. Two carriers of other potentially pathogenic variants in *FANCI* (c.1264G>A; p.G422R and c.1412C>G; p.P471R) also had a family history of OC (**Table 2**). The number of carriers of potentially pathogenic *FANCI* variants with a family history of OC (7/98, 7.1%) was significantly different than isolated cases of HGSC (p=0.027), but there was no significant difference when accounting for family history of OC or BC (p=1). In contrast, *FANCI* c.1813C>T was the only variant identified in 1/63 (1.6%) familial CDN BC cases (**Figure 4c, Supplementary Table 6**) and the carrier is known to be of Greek Canadian origin.

The association of potentially pathogenic variants in HGSC cases was compared with controls in AUS study subjects (**Table 3**). There was no significant difference in allele frequencies of *FANCI* variants in cases versus controls, though odds ratios (ORs) were >1 for three of these variants. As not all *FANCI* variants were identified in AUS controls, we used data from the non-cancer samples in the gnomAD database^61^ to evaluate the ORs. The allele frequency of all gnomAD samples was used as the AUS population is likely of mixed ancestry as exemplified by the observation that c.13A>G; p.I5V was only identified in the East Asian population in gnomAD (**Supplementary Table 6**). The OR was >12 for six of seven of the rarest *FANCI* variants (VAF<0.1%), in contrast to 1.46 for the most common *FANCI* c.1813C>T (**Supplementary Table 8**). *FANCI* c.286G>A had an OR <1 when compared to all gnomAD samples^61^. However, when this analysis was performed with data from the non-Finnish EUR population, the rationale being that over 90% of AUS cases are likely of white EUR ancestry, the OR was >3 (**Supplementary Table 8**). Our findings suggest that rare potentially pathogenic *FANCI* variants, including *FANCI* c.1813C>T, may play a role in hereditary OC cases from AUS cases.

**Table 3.**
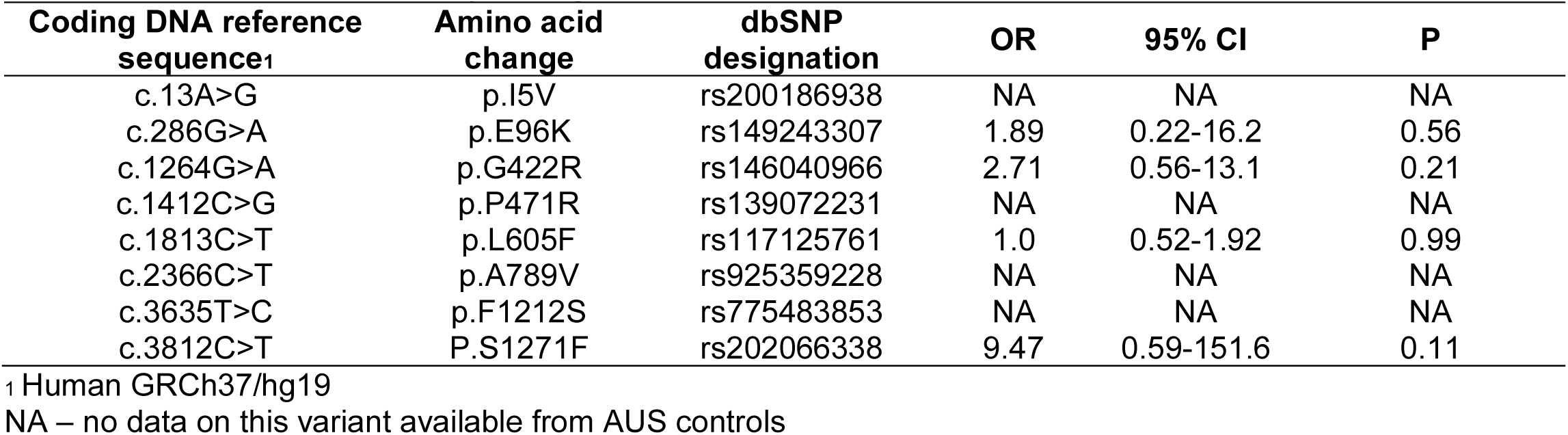
Comparison of potentially pathogenic *FANCI* variant carrier frequencies in AUS HGSC cases to controls.

To further evaluate the effect of *FANCI* variants in conferring risk to OC and BC in non-FC populations, we investigated rare variants (VAF<1%) from imputed SNP array data that was available from two case-control studies: Ovarian Cancer Association Consortium (OCAC)^75^ and Breast Cancer Association Consortium (BCAC)^76,77^. In all, nine missense and one splice site variant were identified in OCAC and BCAC databases. With the exception of *FANCI* c.1813C>T none of the other missense variants were predicted to be potentially pathogenic (**Supplementary Table 9**). The data imputed from the OCAC database^75^ revealed that the OR for *FANCI* c.1813C>T was highest in endometrioid and HGSC subtypes compared to all histopathological subtypes combined, though there was no significant difference in allele frequency in OC cases compared to controls (**Supplementary Table 10**). To compare to a known OC pathogenic variant, OCAC data was investigated for the most common pathogenic *BRCA1* variant reported in FCs, c.4327C>T; p.R1443X, which is one of the most commonly reported variants from populations of EUR descent^46^. The OR was highest in HGSC cases, though there was no significant difference in allele frequency when all OC subtype cases were compared to controls (**Supplementary Table 10**). There was no significant difference in allele frequency between BC cases and controls for *FANCI* c.1813C>T in a similar analysis of BCAC case-control data^76,77^ (*BRCA1* c.4327C>T data was not available in the BCAC database) (**Supplementary Table 10**). These findings show that *FANCI* c.1813C>T carriers occurred in non-FC populations and suggest a trend towards c.1813C>T carriers in HGSC and endometrioid OC as compared to other OC subtypes.

### Clinical features of FC OC from *FANCI* c.1813C>T carriers are similar to those of HGSC cases

We reviewed available clinical characteristics of OC in *FANCI* c.1813C>T carriers as CPGs have been associated with specific subtypes of OC. For example, pathogenic *BRCA1* variants are often associated with HGSC and high-grade endometrioid subtypes of OC and younger age at diagnosis^39,78-81^. Given the paucity of *FANCI* variants, we focused on 13 FC OC carriers of *FANCI* c.1813C>T variant (**Table 1**) and noted that 12 had serous and one had a mucinous subtype OC (**Supplementary Table 7**). The seven carriers found in the context of sporadic OC cases (**Table 1**) were reported as HGSC subtype. The remaining six carriers were identified in OC case with a known family history of cancer (Table 1), where five had serous subtype OC and one had a mucinous subtype OC. Though the sample size was limited, there appeared to be no striking differences in the ages of the diagnosis for OC in carriers where age ranged from 40 to 81 years (average = 59.2 years) as compared with non-carriers in the sporadic OC group (average = 61 years), and aligned with average age of diagnosis of OC in the North American population^1^ (**Supplementary Table 7**).

Survival data, available for sporadic OC cases, showed that all seven *FANCI* c.1813C>T carriers were deceased having had an average survival of 61.1 months (range 9-163). Due to sample size we were unable to perform survival analysis using Kaplan-Meier estimation: 57% (4/5) OC cases did not survive past five years though it should be noted that one of these cases survived 61 months (**Supplementary Table 7**). Survival past 61 months in *FANCI* c.1813C>T carriers (2/7; 28%) is comparable to non-carrier sporadic HGSC cases (100/334; 30%).

### Co-occurrence of other potentially pathogenic variants in OC predisposing genes in *FANCI* c.1813C>T carriers

To further characterize the germline genomic landscape of *FANCI* c.1813C>T carriers, we performed WES analysis of all seven carriers identified in the sporadic FC OC cases and compared it with WES data from five carriers identified in OC families (**Table 1**). We analyzed *FANCI* c.1813C>T OC (n=12) carriers for the co-occurrence of pathogenic variants in known high-risk OC predisposing genes^82^, as these variants are known to significantly increase cancer risk. Only previously known carriers of *BRCA1* (F1055 and F1520) or *BRCA2* (F762) pathogenic variants in familial cases were identified (**Table 1, Supplementary Figure 2**). None of the sporadic OC cases (n=7) carried pathogenic variants in *BRCA1, BRCA2, BRIP1, RAD51C*, and *RAD51D*. Moreover, the *FANCI* c.1813C>T variant did not co-occur in carriers of the recently reported potentially pathogenic *RAD51D* c.620C>T; p.S207L^33^ variant found to recur in sporadic FC HGSC cases^33^.

### FANCI protein is expressed at low-to-moderate levels in HGSC tumour samples

As FANCI protein expression has not been investigated in OC nor fallopian tube epithelial (FTE) cells, a proposed tissue of origin for the HGSC subtype^83-89^, we performed immunohistochemistry (IHC) analysis of an available tissue microarray (TMA) containing cores from formalin-fixed paraffin-embedded (FFPE) tumor tissues. IHC analysis of fallopian tube revealed strong nuclear and low-to-moderate cytoplasmic staining in FTE cells in contrast to stromal cell components where staining was low or undetectable (**Figure 5a, Supplementary Figure 3**). These observations suggest that FANCI protein is robustly expressed in cells associated with the origins of HGSC. In contrast, IHC analysis of tumour cells in HGSC tissue cores exhibited variable staining (**Figure 5b, Supplementary Figure 3**), where the majority (83/94, 88.3%) exhibited low-to-moderate staining intensity. All HGSC tissue cores exhibited strong nuclear and moderate cytoplasmic staining in epithelial cell components, in contrast to the stromal cell components where staining intensity was low or undetectable. A separate IHC analysis of tumour tissues available from eight *FANCI* c.1813C>T carriers revealed a range of staining intensity (**Supplementary Figure 3**), consistent with the expectation that the variant allele could be expressed in tumours (**Figure 2a**). Using Kaplan-Meier survival analysis we found no correlation of staining intensity in epithelial tumour cell components of the HGSC tissue cores with overall or progression free survival (**Figure 5c**). Moreover, age at diagnosis, disease stage, residual disease, chemotherapy type, and survival (disease-free and 5-year) were not correlated with the intensity of protein staining. We were not able to similarly investigate *FANCI* variant c.1813C>T; p.L605F carriers due to the small number of cases. In cases (n=90) that received standard of care chemotherapy (adjuvant taxol and carboplatin) there appeared to be a trend towards lower FANCI staining intensity in HGSC tumour cores from cases having had a prolonged disease-free survival (p = 0.124) (**Figure 5c**).

**Figure 5.**
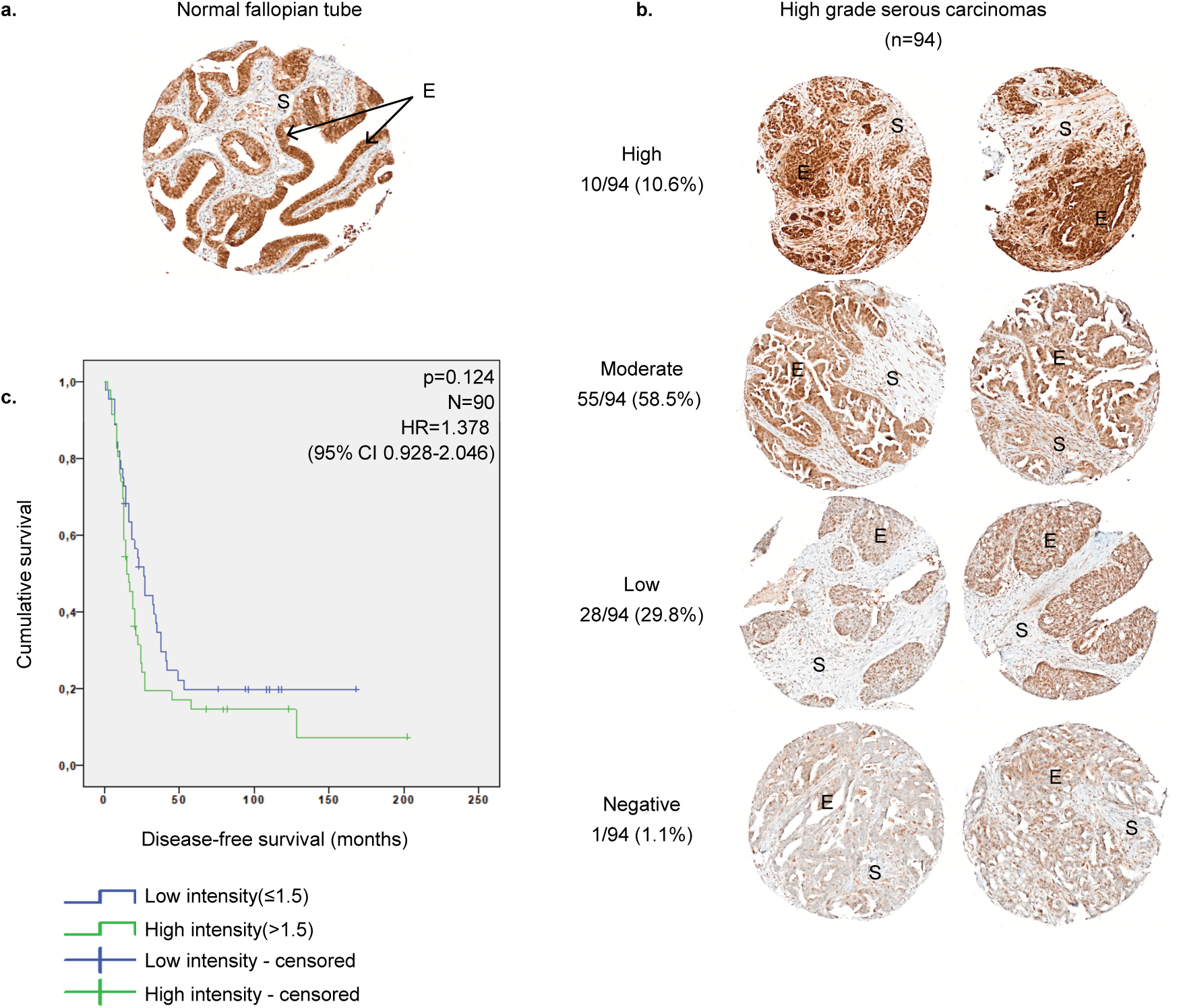
FANCI protein expression in HGSC by IHC of TMA analyses. A) An example of IHC analysis of FANCI protein of a paraffin embedded normal fallopian tube tissue core. B) Examples of different patterns of intensity of IHC analysis of FANCI protein of HGSC tissue cores (epithelial cell components scored). C) Kaplan-Meier survival curve of HGSC cases for disease-free survival (in months) in epithelial cell component. Cases included in the analyses had received only adjuvant taxol and carboplatin chemotherapy. Survival curve was calculated according to Kaplan-Meier method coupled with a log rank test. E=epithelial component; S=stromal component

## Discussion

Our investigation of *FANCI* c.1813 C>T identified by WES analysis of a *BRCA1* and *BRCA2* mutation-negative multi-case OC family of FC descent suggests that this variant may play a role in OC risk. Our investigation was predicated upon the strong founder effects observed in FCs of Quebec and the expectation that candidate risk alleles would recur and thus could be readily identified due to common ancestors in this population^26,27^. Our strategy focused on screening this variant in *BRCA1* and *BRCA2* mutation-negative multi-case OC families of FC descent with the expectation that familial aggregation of OC is not by chance but due to an underlying genetic factor. Given founder effects, we also expected that our candidate alleles would recur in sporadic FC OC cases. Indeed, we showed that *FANCI* c.1813 C>T not only recurs in FCs but is found at a higher frequency in OC cases with a family history of OC relative to sporadic OC cases and cancer-free controls from the same population. Taken together these findings support a role for *FANCI* c.1813C> in modifying OC risk in the FC population.

*FANCI* c.1813C>T was the only potentially pathogenic *FANCI* variant identified in our study of FC cancer cases. Our findings are reminiscent of the recurrence of specific variants in familial FC cancer populations of Quebec, such as *RAD51D* c.620C>T; p.S207L in familial and sporadic OC cases33, PALB2 c.2323C>T; p.Q775X in BC cases and HBC families^32,90^, and MSH6 c.10C>T; p.Q4X in colorectal cancer (Lynch Syndrome) families^91^. Given the genetic demography of the FC population of Quebec, it is likely that carriers of *FANCI* c.1813C>T have common ancestors as has been shown with carriers of specific recurrent pathogenic variants in *BRCA1*^48,92^, *BRCA2*^47,48,93^, and *MSH6*^91^ reported in cancer families. As has been reported with all of these examples of CPGs in non-FC populations, we expected an array of potentially pathogenic variants in our candidate gene. Indeed, we observed eight potentially pathogenic *FANCI* variants in AUS HGSC cases, which included c.1813C>T. A recent genome-wide discovery study of AUS HGSC cases did not report *FANCI* among the list of potentially new CPGs for OC^74^. However, the missense *FANCI* variants found in our analysis of AUS samples, which were investigated in both studies, would have been excluded due to a focus on LoF variants^74^. Our investigation of HGSC cases from the AUS population thus also suggest that FANCI variants may also play a role in OC in non-FC populations.

Although a mechanism of action for *FANCI* in OC risk is unknown, our findings suggest that the *FANCI* c.1813C>T variant affects gene function. This is suggested by the instability of the variant encoded isoform based on our *in vitro* analyses. Though tumour DNA was not available for all of our variant carriers, Sanger sequencing of DNA from FFPE tumour cells suggest loss of the wild type allele and retention of the variant allele had occurred in two FC HGSC *FANCI* c.1813C>T carriers, as shown in **Supplementary Figure 4**. Interestingly, tumour samples from a bilateral OC case predominantly exhibited the *FANCI* variant allele suggesting that loss of the wild type allele could have been be an early event in tumour progression in this case. Also, HGSC samples from both cases had acquired somatic pathogenic variants in *TP53*, a known major driver of the majority of HGSC^94,95^. Our IHC analyses showed differential FANCI protein expression, with a high proportion of HGSC tumour cells exhibiting low-to-moderate levels of protein expression. This is in contrast to robust FANCI protein expression observed in FTE cells, a purported origin of HGSC^83-89^. All of these findings suggest loss of *FANCI* may play a role in OC akin to that suggested by other CPGs in the HR pathway, such as *BRCA1* and *BRCA2*^96^. In keeping with this hypothesis is that loss of the chromosome 15q arm, which contains the *FANCI* locus (15q26.1), has been reported in 55% of 978 HGSC samples by The Cancer Genome Atlas (TCGA) project^95^. A review of available genomic data from TCGA project (cbioportal.org) revealed that five tumour samples likely had homozygous deletions involving the *FANCI* locus and all such samples exhibited low *FANCI* mRNA expression. As expected by observations from previous studies of the genomic landscape of HGSC tumour DNA, somatic potentially pathogenic variants in *FANCI* are also rare (<1%)^95^.

Based on available genetic data from non-Finnish EURs, a population comparable to the ancestral origins of FCs, the allele frequency of *FANCI* c.1813C>T at 1% is higher than expected as compared to many pathogenic variants in established CPGs such as *BRCA1* and *BRCA2*, or the recently proposed OC risk genes, such as RAD51D^33,46^. Similarly, the carrier frequency of c.1813C>T in AUS cancer-free controls at 1.9% was more common than anticipated. The carrier frequency of c.1813C>T in the general population is reminiscent of the more common potentially pathogenic *CHEK2* c.1100delC variant, a moderate risk BC predisposing variant, which was originally identified at a frequency of 1.4% in population controls^97^. Moreover, this *CHEK2* variant was also found more frequently in BC cases from HBC families than sporadic BC cases, relative to healthy controls^97^. Similarly, LoF germline variants in *BRIP1* were more frequently identified in OC cases with a family history of OC than with a family history of BC only, relative to control cohorts^98^. Although our findings of estimating the overall risk to OC using OCAC data was inconclusive, we estimated that carriers of *FANCI* c.1813C>T variant in FC *BRCA1* and *BRCA2* mutation-negative OC families have increased risk based on the OR of 5.6 (p=0.006; 95% CI=1.6-19). Though the confidence interval is wide, due to the small sample size, our findings are supported by the observation that cancer-free *FANCI* c.1813C>T carriers (male or female) were significantly correlated with having a first-degree relative with OC in the FC population, using CARTaGENE data.

It is interesting that *rs8037137*, which is located 1.68 mega-base pairs downstream of *FANCI* c.1813C>T, was among the polymorphic genetic markers found significantly associated with risk to either invasive epithelial or HGSC subtype OC in a large genome wide association analysis of OCAC data^75^. Consistent with these findings is our observation that the OR for *FANCI* c.1813C>T in the OCAC study groups are highest in endometrioid and HGSC subtype OC cases. A similar analysis of other potentially pathogenic *FANCI* variants identified in our study was not possible as corresponding genetic data was not available in the OCAC database. These observations, along with higher frequency of *FANCI* c.1813C>T in familial and sporadic OC cases relative to cancer-free controls, suggest the possibility that *FANCI* c.1813C>T may modify risk to OC at levels lower than high risk pathogenic variants found in *BRCA1* and *BRCA2*. Though the allele frequency of our rare *FANCI* variant was higher in familial OC cases relative to sporadic forms of the disease in FC OC cases, we are mindful of the limitations of our study due to sample size. Therefore to further investigate this hypothesis about levels of risk to OC, we estimated a sample size of approximately 100 OC families and 7,000 female cancer-free controls or 13,000 HGSC cases and 115,000 female cancer-free controls to achieve 80% power would be required (https://www.openepi.com/SampleSize/SSCC.htm), numbers that are currently unattainable in FCs.

The role of *FANCI* c.1813C>T in modifying risk to BC is unclear. Although variant carriers were found in FC familial BC cases, there were proportionally more carriers in BC cases from HBOC families than in HBC families. We also identified a variant carrier in a BC family of Greek Canadian origin: this family was also ascertained from the same catchment area as many of our FC cancer families. In our analysis of FC cancer-free controls that included imputed genotypes, variant carriers were significantly correlated with having a first-degree relative with OC or BC. These findings are in part reminiscent of the variable penetrance for BC and OC for known high risk CPGs, where carriers are more likely to harbour pathogenic variants in *BRCA1* or *BRCA2* (or *PALB2*) based on family history of BC and OC^48^. Thus, it is possible that *FANCI* c.1813C>T confers risk to BC that is lower or more variable than for OC. However, there have been independent reports of BC cases carrying other *FANCI* variants with VAF 10-3 to 10-6 in cancer-free individuals. At least 19 different variants have been described in familial and/or sporadic BC cases: four nonsense, three frameshift, two splicing, and 10 missense variants (see **Supplementary Figure 5** and **Supplementary Table 11**)^99-106^. These *FANCI* variants were reported in Finnish^104^ (4/1524, 0.3%), Chinese^103^ (1/99, 1%), and two Spanish^101,105^ (1/154, 0.6% and 1/94, 1.1%) studies. It is possible that these rare *FANCI* variants, which include putative LoF variants, affect BC risk in various populations.

There have been reports of *FANCI* variant carriers in a variety of cancer types such as prostate cancer^104,107,108^, sarcoma^109^, malignant pleural mesothelioma^110^, acute myeloid leukemia^111^, head and neck carcinoma^112^, and colorectal cancer^113^ (see **Supplementary Figure 5** and **Supplementary Table 12**). Although we also observed colorectal cancers in our *FANCI* variant carrier families of FC descent, a pedigree review found no significant differences in the number of colorectal cancer cases in pedigrees from variant carriers as compared to non-carriers (p=0.47). This observation is consistent with our analysis of family history of cancer-free carriers in the FC population using the CARTaGENE biobank. The role of *FANCI* variants in other cancer types remains to be determined.

Research has shown that FANCI plays a role in the response to DNA damage. FANCI regulates the recruitment of the FA core complex to sites of interstrand crosslinks, and thus plays an important function upstream in the FA-HR DNA repair pathway^114^. FANCI encodes one of only two proteins that comprise the ID2 complex, the other being FANCD2, which is distinct from the FA core complex that includes proteins such as FANCC and FANCM (reviewed by Niraj et al.^115^). *In vitro* cell lines modeling pathogenic variants or gene knock outs of *BRCA1, PALB2*, or *RAD51D* have exhibited sensitivity to cisplatin and PARPi’s, providing some insight into their role in DNA repair^33,116-118^. Our observations in cell lines expressing the FANCI p.L605F isoform differs in that we observed sensitivity to cisplatin but not to the PARPi, olaparib. These findings are consistent with an independent report showing lack of sensitivity to a PARPi (KU0058948) in a fibroblast cell line transduced with HPV E6/E7 from a *FANCI* FA patient, as well as in cell lines generated from *FANCA, FANCL, FANCD2* and *FANCJ* (*BRIP1*) patients^119^. The indirect role of FANCI in HR DNA repair and recent evidence suggesting that FANCI also has functions independent from the FA DNA repair pathway^120-124^ may be consistent with our *in vitro* studies. Two *FANCI* c.1813C>T variant carriers survived over five years after initial diagnosis (nine and 13.5 years). The lengthy survival time is unusual given the overall five- and ten-year survival rates of 32% and 15%, respectively, reported for advanced stage HGSC cases in North America^6^. Further studies of FANCI are warranted as these findings are consistent with our observation of a trend towards low FANCI protein expression with better disease-free survival in HGSC cases, though these findings are not significant.

Although the biological role of FANCI remains to be elucidated, biallelic inactivation of *FANCI* has been associated with FA, a rare disease that is characterized by congenital defects and developmental disabilities^34-36^. FA is a heterogenous disease with 22 known causal genes, where *FANCI* cases comprise approximately 1% of all FA diagnoses^125^. As *FANCI* associated FA cases are rare, the incidence of cancer in biallelic carriers has not been reported. Interestingly, though a Fanci knockout mouse model was recently reported showing a low Mendelian ratio there was no information on cancer incidence^126^.

This is the first study to describe potentially pathogenic variants in *FANCI* in the context of familial OC and in a member of the ID2 complex of the FA DNA repair pathway. Our strategy of investigating familial and sporadic cancer cases from a population showing strong founder effects and pursuing *in vitro* molecular analysis of missense variant found to recur in cancer cases contributed to the discovery of *FANCI* as a new candidate OC risk gene. While the feasibility of experimenting with missense variants at the molecular level can be an issue, this study reveals the importance of pursuing such variants during the gene discovery phase, especially when plausible candidates based on gene function are revealed by WES analyses of defined cancer families. Although some of the identified *FANCI* variants are predicted to affect gene function as shown in our *in vitro* analyses of *FANCI* c.1813C>T encoded isoform, further studies are warranted to evaluate their role in OC risk. Establishing risk is important in the context of familial aggregations of OC and host behaviours that modify risk to OC, as has been shown with OCP use in carriers of pathogenic *BRCA1* and *BRCA2* variants^70^. An investigation of carriers of potentially pathogenic *FANCI* variants is also warranted given the intriguing observation of sensitivity to cisplatin but not to olaparib in our *in vitro* studies of *FANCI* c.1813C>T, as this might impact the efficacy of PARPi’s in the treatment of HGSC in these cases.

## Methods

### Study subjects, samples, and DNA sequencing

Information about the study subjects, PBL and tumour DNA samples, and FFPE tissues of HGSC cases and normal fallopian tube used for TMAs were obtained from various biobanking resources (**Supplementary Table 1**). The OC samples from Réseau de recherche sur le cancer (RRCancer) Tumour and Data biobank derived its collection from patients attending a major gynecologic oncology hospital centre in Montreal. This centre largely services FCs, where it is estimated that 85% of samples come from participants who self-identify as FC^46^. Samples within this collection with a familial history of OC and/or BC have been extensively studied, where the majority self-report grandparental FC ancestry of index cancer affected cases^30,48,49^. This project has received approval from The McGill University Health Centre (MUHC) REB (MP-37-2019-4783 and 2017-2722). To further protect the anonymity of study subjects, all samples were assigned a unique identifier.

#### Discovery of FANCI c. 1813C>T in an OC family

The *FANCI* c.1813 C>T variant was initially discovered by WES and bioinformatic analyses of PBL DNA from sisters, both with OC, from family F1528 that were performed at the McGill Genome Centre (MGC). This multi-case OC family of FC descent, available from the RRCancer biobank, has been described previously as a *BRCA1* and *BRCA2* mutation-negative family^39^ and has since been updated to include new information, including histopathology of OC and a reported case of ear, nose, and throat cancer (**Figure 1**). PBL DNA (~500 ng) from two sisters from this family was captured with the Agilent SureSelect 50 Mb exome capture oligonucleotide library, and then sequenced with paired-end 100 bp reads on Illumina HiSeq 2000. After removing putative PCR-generated duplicate reads using Picard (V.1.48), sequencing reads were aligned to human genome assembly hg19 using a Burroughs–Wheeler algorithm (BWA V.0.5.9). Sequence variants were called using Samtools (V.0.1.17) mpileup and varFilter meeting the following criteria: at least three variant reads, ≥20% variant reads for each called position, and Phred-like quality scores of ≥20 for SNPs and ≥50 for small insertions or deletions. Annovar^127^ and custom scripts were used to annotate variants according to the type of variant, Single Nucleotide Polymorphism database designation (dbSNP), Sorting Intolerant from Tolerant (SIFT) score^128^, and allele frequency data from the 1000 Genomes Project^45^ and NHLBI ESP v.2014 (https://evs.gs.washington.edu/EVS/). Then, the variant list was organized to select top candidate potentially pathogenic variants that were shared in common among the two sisters by de-prioritizing the following: (1) synonymous or intronic variants other than those affecting the consensus splice sites; (2) variants seen in more than 5 of 416 exomes from patients with rare, monogenic diseases unrelated to cancer that were independently sequenced and available at the MGC and (3) variants with a frequency ≥1% in either the 1000 Genomes Project or NHLBI exome datasets. Using a candidate gene approach, we then further prioritized the list of candidates based on their role in FA-HR pathways. Using this strategy, *FANCI* c.1813C>T was the only candidate remaining on the list of prioritized variants (n=276) shared in common between the two sisters in family F1528. The presence of the *FANCI* variant was verified using Integrative Genomics Viewer (IGV)^129^. The *FANCI* c.1813C>T variant was validated by targeted PCR analysis and Sanger sequencing at the MGC using standard methods (see Supplementary Table 13).

Since the initial discovery of the *FANCI* variant in family F1528, newer WES technology and bioinformatic tools became available, and thus we repeated our analysis with DNA from the same sisters from this family. WES and bioinformatic analyses were again performed at the MGC using Roche NimbleGen SeqCap® EZ Exome Kit v3.0 (Roche Sequencing) followed by HiSeq 100 bp paired-end sequencing (Illumina) applying manufacturer’s protocols. Sequencing reads were aligned to human genome assembly hg19 using BWA-MEM v0.7.17, then deduplicated using Picard v2.9.0 (Broad Institute). Bases were recalibrated using the GATK best practices. Variants were called using HaplotypeCaller available from GATK v3.5 (Broad Institute) and recalibrated according to GATK best practices. The filtered variants were then annotated and loaded into a GEMINI v0.19.1 database as per the recommended workflow. Data was filtered for non-synonymous rare variants (VAF<1%) deduced from a publicly available database gnomAD v2.1.1^61^ identified in genes with reported function in DNA repair pathways (n=276^130^). *FANCI* c.1813C>T was once again the only variant directly involved in the FA-HR DNA repair pathway identified in both sisters. The presence of the *FANCI* variant was again confirmed by IGV^129^ and validated by PCR analysis and Sanger sequencing at the MGC using standard methods (see **Supplementary Table 13**).

#### Genetic analysis of FANCI c. 1813C>T in FC cancer cases

The allele frequency of *FANCI* c.1813C>T was determined by investigating selected index OC or BC cases defined based on family history of OC and/or BC or sporadic disease where cases were not selected based on family history of cancer, where all were-self-reported FC descent as previously described^30,48,49^ (see **Supplementary Table 1**). The average age of diagnosis for OC families was 49.8 years (range 24-77). The average age of diagnosis was 44.6 years (range 22-65) and 43.7 years (range 18-65) for HBC and HBOC families, respectively. Sporadic BC cases were diagnosed with invasive BC before the age of 70 (average = 52.7, range 25-69)^131^. We cannot exclude the possibility that some cases occurred in more than one study group: based on RRCancer biobanking sample number, OC cases from at least 13 families were also found in pedigrees from BC cases that were genotyped from the familial HBOC study group.

Carriers of *FANCI* c.1813C>T were identified by targeted genotyping of PBL DNA samples or from surveying available WES data (subjected to the same latest WES technology and data analysis pipeline as described above) from affected cases in our study groups (see **Supplementary Table 1**). PBL DNA from OC or BC cases were genotyped using a custom TaqMan® genotyping assays based on established methods (see **Supplementary Table 14**). Where normal DNA was no longer available from the study case, genomic DNA extracted from the tumor (if available) was provided by the RRCancer biobank for genotyping. PBL DNA from sporadic BC cases were genotyped using Sequenom® iPLEX® Gold Technology at the MGC^131^. Samples that were removed from the analysis were due to poor DNA quality (n=30), duplication (n=1), or were from cases exceeding age limit criteria (70 years or older when diagnosed with first invasive BC; n=2). Results from a total of 558 cases were evaluated for *FANCI* c.1813C>T carrier status. The *FANCI* locus (NC_000015.9: g.89828441C>T) was reviewed in WES data, validated by IGV analysis, and *FANCI* c.1813C>T variant carriers verified by Sanger sequencing as described (see **Supplementary Table 13**).

#### Genetic analysis of FANCI c. 1813C>T in FC cancer-free controls

Genotyping data was obtained from CARTaGENE (www.cartagene.qc.ca) from cancer-free FC subjects as controls. The overall average age in this study group was 54.45 years (range 39.06-71.05)^60^. The study subjects were comprised of 2,627 females (average age = 54.45 years; range 39.06-71.05) and 2018 males (average age = 55.3 years; range 39.1-70.9). Selection criteria for individuals with genotyping data are biased towards individuals with higher quantity of health data (see **Supplementary Table 4**). Individuals were defined as FC if they were born in Quebec, their parents and all four grandparents were born in Canada, and French was the first language learned. Genotyping data was available from samples that were genotyped in three different batches of experiments that used two different genotyping platforms (Illumina and Affymetrix; see **Supplementary Table 4**). Data was imputed when there was no representative probe for a locus on the genotyping array using the Sanger Imputation Service (https://imputation.sanger.ac.uk/) with Haplotype Reference Consortium (release 1.1) as the reference panels. Pre-phasing and imputation was performed using Eagle2^134^ and the positional Burrows-Wheeler transform (PBWT)^135^. Samples were removed as part of quality control to improve imputation of the array (see **Supplementary Table 4**).

#### Statistical analyses of FANCI c. 1813C>T frequency in FC cancer cases and controls

Two-sided Fisher exact test was used to compare frequencies of *FANCI* c.1813C>T carriers in the cases and controls or between different study subjects, where a p-value of 0.05 was considered significant. ORs and 95% confidence intervals were estimated for all study subjects for this allele.

#### Genetic analysis of FANCI locus in FC cancer cases

The *FANCI* locus was investigated in available WES data from 157 OC or BC cases of FC descent (see **Supplementary Table 1**). Rare (VAF<1%) *FANCI* variants identified in WES data were further subjected to bioinformatic analyses using up to 16 *in silico* predictive of the effect of the nucleotide change(s), which includes two tools to predict splice site variants. Variants were considered potentially pathogenic if they were predicted to be pathogenic/deleterious in ≥8 tools for missense variants or two tools for splice site variants. Nonsense variants were considered potentially pathogenic but in-frame deletions were not. Variants were annotated using the Ensembl Variant Effect Predictor^136^.

#### Genetic analysis of FANCI c. 1813C>T in OC and BC cases and controls from consortia databases

The *FANCI* locus was investigated in available OCAC and BCAC data. Summary statistics including log_2_OR, standard error (SE), X_2_, and p-value for 25,509 epithelial OC cases (22,406 invasive) and 40,491 controls of EUR ancestry^75^ were obtained from OCAC (http://ocac.ccge.medschl.cam.ac.uk). Summary statistics were available for histopathological subtypes as well as for the entire cohort in OCAC and have been previously summarized^75^. Summary statistics including log_2_OR, standard error (SE), X_2_, and p-value for 46,785 cases and 42,892 controls of EUR ancestry^76,77^ were obtained from BCAC (http://bcac.ccge.medschl.cam.ac.uk). Details of the subjects and genotyping analyses is described elsewhere^75-77^. Summary statistics for *BRCA1* c.4327C>T and *rs8037137* loci were obtained as comparators (see **Supplementary Table 10**). All rare (VAF<1%) *FANCI* variants identified in OCAC and BCAC were subjected to the same bioinformatic analyses using *in silico* tools as described.

#### Genetic analysis of FANCI locus in AUS HGSC cases and controls

The *FANCI* locus was investigated in germline sequencing data available from WES analysis of HGSC cases ascertained from the AUS population as previously described^74^. Briefly, all AUS cases had ovarian, fallopian tube, or peritoneal cancer (n=516) and did not carry pathogenic variants in *BRCA1* and/or *BRCA2* (see **Supplementary Table 1**). AUS controls were ascertained from the lifepool study as previously described^137^. Cancer-free women with available sequencing results for *FANCI* (n=4,878) were included in this study. The identified rare (VAF<1%) variants found in *FANCI* were subjected to the same bioinformatic analyses using *in silico* tools as described.

#### Genetic analysis of FANCI locus in CDN BC cases

The *FANCI* locus was investigated in germline sequencing data available from WES analysis of PBL DNA from female subjects with OC, BC, or pancreatic cancer (n=63) who were recruited from centres in Montreal via the McGill Cancer Genetics Programme. All recruited individuals had a strong family history of BC. A BRCAPro score^138^, which is based on studies of Ashkenazi Jewish and EUR descent individuals, was generated to predict the likelihood of families carrying pathogenic variants in *BRCA1* or *BRCA2*. Individuals with a BRCAPro score of >10%, but with no pathogenic variants in these genes were selected. Of this set, 14 individuals were of Ashkenazi Jewish ancestry. *FANCI* variants were selected from PE125 WES data that was generated using the Nextera Rapid Capture Exome enrichment kit (Illumina) followed by HiSeq-4000 sequencing performed by the CRUK CI genomics core facility in the UK. Variant Call Format files were generated with a standard pipeline following GATK Best Practices recommendations for WES data. The identified rare (VAF<1%) variants found in *FANCI* were subjected to the same bioinformatic analyses using *in silico* tools as described.

#### Genetic analysis of variants in known OC predisposing genes and DNA repair genes in FANCI c. 1813C>T carriers

Rare (VAF<1%) variants that were identified in known high risk epithelial OC predisposing genes in the analysis of WES data from *FANCI* c.1813C>T carriers was investigated using various bioinformatic tools. Pathogenicity was evaluated in BRCA Exchange^31^ for *BRCA1* and *BRCA2* variants and ClinVar (https://www.ncbi.nlm.nih.gov/clinvar/) for all variants. Rare (VAF<1%) variants in DNA repair pathway genes (n=276^130^) were evaluated in *FANCI* c.1813C>T carriers. The only variant identified that was shared in all cases was *POLG* c.2492A>G and it was pursued further as described below.

The allele frequency of *POLG* c.2492A>G was determined by investigating selected index OC or BC FC cases as above. Carriers of *POLG* c.2492A>G were identified by targeted genotyping of PBL DNA samples or from surveying available WES data from affected cases from our study groups as described. PBL DNA from OC or BC cases were genotyped using a custom TaqMan® genotyping assays based on established methods (see **Supplementary Table 14**). *POLG* c.2492A>G was reviewed in available WES data as above. Genotyping data from CARTaGENE for cancer-free FC controls was investigated as above, including imputation (see **Supplementary Table 4**). *POLG* c.2492A>G was subjected to the same bioinformatic analyses using *in silico* tools as described.

### In vitro analysis of *FANCI* c.1813C>T variant and FANCI encoded protein

#### Cell lines, cell culture and reagents

HeLa cells were grown in Dulbecco’s modified Eagle’s medium (Corning™ cellgro™), supplemented with 10% Fetal Bovine Serum (Gibco™), at 37°C, 5% CO^2^, 20% O^2^. HeLa cells KO for *FANCI* were obtained using the ALT-R CRISPR-Cas9 system from Integrated DNA TechnologiesTM. Cells were transfected with crRNA:tracrRNA:Cas9 RNP-complexes (crRNA sequence: AATCCCCCGATTCCACCAAC), according the manufacturer’s guidelines for RiboNucleoProtein transfection using RNAimax. After transfection, genomic DNA from the pool of transfected cells was extracted using QIAamp DNA Mini Kit (Qiagen, ref 51306). A 500 bp DNA region containing the sgRNA complementing sequence was amplified by PCR from 400 ng of genomic DNA with the Thermo Scientific™ Phusion™ High-Fidelity DNA Polymerase and verified by sequencing using the following primers: Forward: 5’-GTTACTGGACTTCTCAAAAGCTGTAAG-3’ and Reverse: 5’-CTAGGTTGGGCACTTAAGTTTTCCT-3’. Sequencing results from non-transfected cells and genetically altered cells were compared using TIDE software to estimate the percentage of genetically altered cells. Clones were then generated and selected based on FANCI protein depletion using Western blot analysis. Two clones, clones 1 and 2, were used in this study.

When specified, cells were treated with MMC from *Streptomyces caespitosus* (Millipore-Sigma, ref M0440) or Formaldehyde (BAKER ANALYZED® ACS, J.T. Baker®, ref CAJT2106). For protein stability assays, CHX (Millipore-Sigma, ref C4859) was used at a final concentration of 100 μg/ml.

#### siRNA transfection and complementation assays

Approximately 2.5×10_5_ HeLa cells were transfected with 50 nM of siCTL (UUCGAACGUGUCACGUCAA) or siFANCI (UGGCUAAUCACCAAGCUUAA) with RNAimax (Invitrogen) according the manufacturer’s protocol. Then, after 24 hours, cells were transfected again with the same siRNAs. After six hours, cells were complemented with the indicated constructs of Flag-FANCI or pcDNA3 plasmid (empty vector) using Lipofectamine 2000 according to the manufacturer’s protocol, using the following quantities of plasmids: 1 µg of WT, 3 µg of L605F and 1.5 µg of P55L.

#### Protein extraction and immunoblotting

Cells were collected by trypsinization and rinsed once in cold PBS. Cell pellets were then incubated in lysis buffer (10 mM HEPES pH 7.4, 10 mM KCl, 1% Triton, 150 mM NaCl, 30 mM Na_2_P_2_O_7_.10H_2_O, 1 mM EDTA and 1 µg/ml Leupeptin, 3.4 µg/ml Aprotinin, 1% PMSF, 5 mM NaF, 1 mM Na_3_VO_4_, Complete™ EDTA-free Protease Inhibitor Cocktail [Roche]) for 30 minutes on ice. Cell lysates were then sonicated for 5 minutes (30 seconds on, 30 seconds off, high, Bioruptor) and centrifuged for 30 minutes, 13000 rpm, 4°C. Supernatant was then processed for immunoblotting analysis using the indicated antibodies.

#### Antibodies for Western blotting and immunofluorescence assays

The antibodies used were anti-FANCI (A7) (Santa Cruz Biotechnology, ref sc-271316, 1:100), anti-FANCD2 (Novus, ref NB100-182D1, 1:5000 for western-blot, 1:1000 for immunofluorescence), and anti-vinculin (Sigma, ref V9131, 1:100000). Horseradish peroxidase-conjugated anti-rabbit IgG or anti-mouse (1:10000; Jackson ImmunoResearch) were used as secondary antibodies.

#### Cisplatin and olaparib cell survival assays

Approximately 3×10_5_ HeLa Fanci^-/-^ cells were seeded into one well of a six-well plate. After 24 hours, cells were complemented with the indicated Flag-FANCI construct using Lipofectamine 2000 (Invitrogen), and then after another 24 hours seeded in triplicate into a Corning 3603 black-sided clear bottom 96-well microplate at a density of 3500 cells per well. The remaining cells were stored at −80°C until processing for protein extraction and immunoblotting as described above. Once attached to the plate, the cells were exposed to different concentrations of either 0-300 nM cisplatin (Tocris, #2251) or 0-2.5 μM olaparib. After three days of treatment, nuclei were stained with Hoechst 33342 (Invitrogen) at 10 µg/ml in media for 45 minutes at 37°C. Images of entire wells were captured at 4× magnification using a CytationTM 5 Cell Imaging Multi-Mode Reader and Hoechst-stained nuclei were quantified with the Gen5 Data Analysis Software v3.03 (BioTek Instruments). Cell viability was expressed as percentage of cell survival in olaparib-treated cells relative to vehicle (DMSO)-treated cells. Results represent the mean ±SEM of at least three independent experiments, each performed in triplicate.

#### Immunofluorescence analyses

HeLa Fanci^-/-^ cells were complemented with FLAG-FANCI variants and 0.1 µg of peGFP (when specified) as described above. HeLa Fanci^+/+^ cells were transfected with siRNA and complemented with siRNA resistant FANCI variants as described above. After 18 hours, cells were seeded on a glass coverslip for eight hours and then treated with 50 ng/ml MMC for 18 hours and processed for immunofluorescence with anti-Flag (Cell signaling Technologies, ref 8146, 1:1600) and anti-FANCD2 (Novus, ref NB100-182D1, 1:800) antibodies according to the protocol provided by Cell signaling technologies for Flag antibody. Alexa Fluor secondary antibodies from Life Technologies (ref A-11001, A-11004, A21235) were used at 1:1000. In HeLa Fanci^-/-^ cells, FANCD2 foci were counted in GFP positive cells. In siRNA transfected cells, Flag positive cells were taken into consideration for the quantification of FANCD2 foci. Statistical analysis was performed using GraphPad Prism 8. Mean with SEM are shown, and a Kruskall-Wallis test was performed.

#### Anti-Flag pulldown assays

After siRNA transfection and complementation with Flag-FANCI WT or L605F, HeLa FANCI^+/+^ cells were lysed in lysis buffer (50 mM Tris–HCl, pH 7.5, 150 mM NaCl, 0.5% NP-40) containing protease and phosphatase inhibitors (1 mM PMSF, 0.019TI U/ml aprotinin, 1 mg/ml leupeptin, 5 mM NaF, and 1mM Na_3_VO_4_) incubated for 30 minutes on ice, and lysed by sonication. Insoluble material was removed by high-speed centrifugation (13000 rpm at 4°C) and each immunoprecipitation (IP) was carried out using soluble protein extract in 1 ml of lysis buffer. Fifty ml of anti-Flag M2 affinity gel (Sigma) and 70 U of DNase I were added and incubated at 4°C for 2.5 hours. Beads were washed three times with washing buffer (50 mM Tris–HCl pH 7.5, 250 mM NaCl, 0.5% NP-40) and proteins were eluted with 60 µl of Laemmli buffer. Proteins were visualized by western blotting using the appropriate antibodies.

### Differential FANCI protein expression by IHC analysis of HGSC tumours and normal tissues

Slides containing 4 micron slices of TMAs containing 0.6 mm FFPE tissue cores (spaced 0.2 mm apart) of HGSC (n=101)^139^ and normal fallopian tube (n=15) tissues, and *FANCI* c.1813C>T carrier tumour tissues (n=8) were stained using the BenchMark XT automated stainer (Ventana Medical System Inc., Roche). Antigen retrieval was carried out with Cell Conditioning 1 solution for one hour. The FANCI polyclonal antibody (Sigma HPA039972 dilution 1/200) was automatically dispensed and the TMAs were incubated at 37°C for one hour. The Ultra View DAB detection kit was used, and the slide was counterstained with hematoxylin. The TMAs were scanned with a 20× 0.75 NA objective by VS-110 Olympus.

Staining patterns were evaluated by two independent observers. Intensity of staining was scored for all cores using a four-point system; zero referring to no detectable staining to three referring to the highest staining intensity. As each sample was present in the TMA in duplicate, each case received four scores (two for the first core and two for the second core). The mode score was used for analysis where possible, otherwise the average score was used. The interobserver correlation for IHC analysis of the TMA of HGSC samples was 89%. Staining patterns and analyses from the TMA containing HGSC samples and normal fallopian tube samples were evaluated without prior knowledge of carrier status for *FANCI* c.1813C>T. All HGSC and normal fallopian tube samples were genotyped for *FANCI* c.1813C>T variant as described, and one previously known carrier was identified (PT0004). Samples that could not be scored were removed from further analysis (n=7 HGSC samples, n=2 FTE samples). A second TMA that contained 10 samples from eight *FANCI* c.1813C>T carriers (in duplicate) were also scored separately: the results from one sample from this TMA was removed from analysis due to poor tissue quality.

Spearman correlation was used to measure the strength of the correlation of staining intensity and survival data with clinical data as continuous variables. Survival curve was calculated according to Kaplan-Meier method coupled with a log rank test. Univariable Cox hazard models were used to estimate the hazard ratio as categorical data. All statistical analysis was done using Statistical Package for the Social Sciences software version 24., (SPSS, Inc) and results deemed statistically significance at p<0.05.

## Data Availability

Unknown

## Acknowledgements

We thank Yan Coulombe, Dr. Amélie Rodrigue, and Dr. Faezeh Vasheghani Farahani for technical support, Dr. Lili Fu for reviewing pathology reports, Daryl A. Ronato for graphical support, and the Molecular Pathology core facility of the Centre de recherche du Centre hospitalier de l’Université de Montréal (CRCHUM) for performing the immunohistochemistry. C.T.F. was supported by The Research Institute of the McGill University Health Centre Studentship (RI-MUHC) Award and Ovarian Cancer Canada Trainee Travel (Research) Award. W.M.A. is supported by a Taibah University Scholarship Award and The Ministry of Higher Education, Saudi Arabia. H.F. was supported in part by the Michèle St-Pierre and Canderel fellowships of the Institut du cancer de Montréal. L.G.-S. is supported by a Fonds de recherche du Québec-Santé (FRQS) Fellowship Award and J.-Y.M. is a FRQS Research Chair in Genome Stability. The RI-MUHC and CRCHUM receive support from the FRQS. Research was supported by the following: The Canadian Institute for Health Research (CIHR) operating grants [PCC-156736 to P.N.T., C.M.T.G., J.R.; PJT-156124 to P.N.T., J.R.]; Cancer Research Society and Ovarian Cancer Canada partnership grant [21123 to P.N.T.; Department of Medicine, McGill University Grant to P.N.T.; FRQS and Quebec Breast Cancer Foundation network grants to P.N.T.; Compute Canada resource allocation project wst-164 and Genome Canada Genome Technology Platform award to J.R.; CIHR foundation grant to J.-Y.M. Ovarian tumor banking was supported by the Banque de tissus et de données of the Réseau de recherche sur le cancer of the FRQS affiliated with the Canadian Tumor Repository Network (CTRNet). Funding for BCAC and iCOGS came from: Cancer Research UK [grant numbers C1287/A16563, C1287/A10118, C1287/A10710, C12292/A11174, C1281/A12014, C5047/A8384, C5047/A15007, C5047/A10692, C8197/A16565], the European Union’s Horizon 2020 Research and Innovation Programme (grant numbers 634935 and 633784 for BRIDGES and B-CAST respectively), the European Community’s Seventh Framework Programme under grant agreement n° 223175 [HEALTHF2-2009-223175] (COGS), the National Institutes of Health [CA128978] and Post-Cancer GWAS initiative [1U19 CA148537, 1U19 CA148065-01 (DRIVE) and 1U19 CA148112 - the GAME-ON initiative], the Department of Defence [W81XWH-10-1-0341], and the Canadian Institutes of Health Research CIHR) for the CIHR Team in Familial Risks of Breast Cancer [grant PSR-SIIRI-701]. All studies and funders as listed in Michailidou K et al (2013 and 2015) and in Guo Q et al (2015) are acknowledged for their contributions.

## Author contributions

C.T.F. performed targeted genetic experiments, bioinformatic and statistical analyses, wrote and edited initial drafts of the manuscript. L.G.-S. performed *in vitro* experiments and edited drafts of the manuscript. W.M.A aided in the bioinformatic analyses. T.R. and J.N. performed bioinformatic analyses and edited draft. K.K.O. performed genetic analyses. K.B. aided in experiments. S.L. A. aided in and oversaw experiments. S.B. performed targeted genotyping experiments. C.S. aided in study sample and clinical data collection. L.M. and H.F. performed immunohistochemistry of tissue microarray and associated statistical analyses. E.F., D.N.S., Z.E., and D.P. provided study samples, clinical data, and critical input. W.D.F and A.-M.M.-M. provided study samples and clinical data, critical input, and edited draft. J.M. oversaw whole exome sequencing and bioinformatic analyses of discovery family and edited draft. M.T., P.A.J., and I.G.C. provided genotyping and clinical data, critical input, and edited draft. C.M.T.G. oversaw bioinformatic analyses. J.R. oversaw processing of whole exome sequencing data, developed data analysis pipelines, and oversaw bioinformatic analyses. J.-Y.M. oversaw in vitro experiments. P.N.T. conceived and designed the overall project, wrote the manuscript, and oversaw all aspects of the project. All authors reviewed and edited the manuscript.

